# Changes in inter-limb coordination and kinetics due to gradually introduced locomotor adaptation in people with trans-tibial amputation

**DOI:** 10.1101/2025.07.18.25331783

**Authors:** Brian P. Selgrade, Young-Hui Chang

## Abstract

People with amputation walk asymmetrically, leading to increased risk of intact leg injury. Split-belt treadmill walking using error augmentation has potential to correct this asymmetry. The purpose of this study was to assess how people with trans-tibial amputation and matched control subjects would adapt their limb forces to gradual onset split-belt treadmill walking. Consistent with prior split-belt results in sudden onset split-belt walking, we hypothesized that, after gradual onset split-belt walking, people with trans-tibial amputation and intact controls would display aftereffects in braking force but not propulsive force. We also hypothesized that both groups would have aftereffects in step length symmetry and double support, indicating predictive control of inter-leg coordination. People with trans-tibial amputation and control subjects displayed aftereffects in braking force, propulsive force, double support time, and step length symmetry. People with trans-tibial amputation displayed an aftereffect in step length opposite their baseline asymmetry. Both subject groups had aftereffects in fast (intact) leg forces that were larger for braking and smaller for propulsive forces than baseline. These findings indicate that gradual onset split-belt adaptation involves predictive control of inter-leg coordination and leg forces, which is not impaired by trans-tibial amputation. Predictive control of step length and braking is consistent with prior work, but these results suggest different adaptive control of propulsion than prior sudden onset research. This study shows that gradual onset split-belt walking may correct step length asymmetries in people with trans-tibial amputation, but increased intact leg braking aftereffects has potentially negative implications for correcting amputation-related kinetic asymmetries.

## Introduction

Approximately 1.6 million Americans lived with amputation in 2005, and this population is estimated to increase to 3.6 million by 2050 (Ziegler-Graham et al. 2008). Trans-tibial amputation was the second most common type of amputation in the United States, accounting for over 270,000 amputations between 1988 and 1996 alone (Dillingham et al., 2002). People with trans-tibial amputation create larger forces with their intact leg than with a passive, prosthetic leg (Sanderson and Martin, 1997, Adamczyk and Kuo, 2015, Snyder, et al. 1995), which typically leads them to take longer steps with their prosthetic leg leading than with their intact leg leading (Isakov et al., 1997). They also spend more time in stance on their intact leg (Breakey, 1976) and exert more force on the ground with their intact leg (Baker and Hewison, 1990). These asymmetries illustrate a general overuse of the intact leg during gait, which may relate to higher incidences of osteoarthritis in the intact leg driven by reduced prosthetic side propulsion (Morgenroth et al., 2014, Morgenroth et al., 2011).

Because clinical settings are rarely conducive to measuring ground reaction forces, improving kinematic asymmetry is a commonly assumed clinical goal (Childers and Kogler, 2014). Specifically, several studies have attempted to use split-belt treadmill walking to correct baseline asymmetries, including in stroke survivors (Reisman et al. 2007, Tyrell et al. 2015) and Parkinson’s disease patients (Roemmich et al, 2014). These studies follow the principle of error augmentation, in which the split-belt condition exacerbates the clinical population’s baseline error (i.e. step length asymmetry). When subjects adapt to the split-belt condition, they correct this error over time. Upon removal of the split-belt condition, an aftereffect in the opposite direction of the initial asymmetry occurs. If the leg that leads when subjects take longer steps in baseline is placed on the slow belt during the split-belt condition, the result can be aftereffects that are more symmetric than in baseline or asymmetric in the direction opposite to baseline asymmetry (Reisman et al. 2007, Tyrell et al. 2015). Although this more symmetric aftereffect washes out after one exposure to split-belt treadmill walking, multiple exposures can lead to a more permanent, newly learned motor pattern (Reisman, et al. 2010). For example, among a group of stroke survivors who performed split-belt treadmill walking 3 times per week for 4 weeks, those with the largest baseline step length asymmetries showed reductions in step length asymmetry that were sustained 3 months later (Reisman et al. 2013). Essentially, the aftereffect was no longer washing out. Therefore, the aftereffects of adaptation can provide a glimpse of what may occur in longer-term motor learning but can be studied in a more tractable, controlled manner (Reisman, et al. 2010).

More recently, Darter and colleagues used this error augmentation paradigm to study split-belt walking adaptation in people with trans-tibial amputation (2017). They found that people with traumatic, trans-tibial amputation behaved similarly to control subjects, adapting step length and exhibiting an aftereffect in step length when the belts returned to moving at the same speed (tied belt condition). In another error augmentation study, people with non-traumatic, trans-tibial amputation also adapt their step length to split-belt walking and display aftereffects in step length when returning to tied-belt walking (Kline et al., 2019). These studies extended previous results (Selgrade, et al., 2017) by showing that people with trans-tibial amputation would adapt step length and demonstrate after effects regardless of which foot was on the fast belt. The ability to adapt step length despite the loss of distal motor function and sensory feedback suggests that an error augmentation paradigm could improve symmetry in people with trans-tibial amputation. Both of these error augmentation studies sought to use split-belt adaptation to correct spatial asymmetries (step length asymmetry) in people with trans-tibial amputation. In such studies, the goal is to correct the baseline step length asymmetry in the aftereffect. Such a change from baseline in the aftereffect indicates predictive control of a parameter, in which a central motor program changes the parameter over many strides in response to a change in the environment. Alternatively, reactive accommodation occurs when the parameter is changed immediately due to feedback from the same stride.

Whereas most prior split-belt studies have investigated kinematic asymmetries, fewer have examined kinetic changes in split-belt walking at the limb level (Roemmich et al., 2012, Mawase et al. 2013, Roemmich et al, 2014, Selgrade et al. 2017, Sanchez et al., 2017), and none have assessed kinetic responses to gradual onset split-belt walking. Furthermore, despite the recommendation that forces should be measured during split-belt walking studies (Scarano et al., 2021), the effects of split-belt walking on ground reaction forces in people with amputation are unknown. People with trans-tibial amputation generally produce lower force from the prosthetic leg, because a passive prosthetic ankle produces much less power than a biological ankle (Bateni and Olney, 2002, Zmitrewicz et al., 2007). However, in order to increase intact leg leading step length, we would expect people with trans-tibial amputation to increase work from their amputated leg. Since this work is unlikely to come from a passive prosthetic ankle, it may come during single limb support from the hip on the amputated side, which generally does more work than a healthy individual’s hip in normal walking (Silverman, et al., 2008, Adamczyk and Kuo, 2015). Work from the hip in single support is less efficient at propelling a person forward than work from the trailing ankle just before contralateral heel strike (Kuo, 2002). These dynamics suggest that an error augmentation paradigm that corrects baseline step length asymmetry in the aftereffect would be have the most potential benefit to people with amputation if it also induced more propulsion from the prosthetic side, which would be indicated by a higher propulsive force from the prosthetic leg in the aftereffect).

The purpose of this study was to assess adaptation of leg forces in people with amputation during split-belt treadmill walking. Our primary hypothesis was that, after undergoing gradual onset split-belt walking, both people with trans-tibial amputation and intact subjects would display aftereffects in braking GRF but not propulsive GRF. This result would suggest that, just as in healthy individuals (Ogawa et al., 2014), braking is controlled predictively while propulsion is controlled reactively in people with adaptation. Lastly, we hypothesized that, consistent with prior studies, people with trans-tibial amputation and intact subjects would both display aftereffects in step length symmetry and double support time upon reintroduction of the tied belt condition. Building upon previous results finding predictive control of step length adaptation in amputees (Darter et al. 2017, Kline et al., 2019, Selgrade et al., 2017) and double support time in healthy individuals (Reisman et al., 2005), results supporting our last hypothesis would indicate that predictive control of interlimb coordination in people with amputation extends to temporal variables.

## Methods

### Subjects

Sixteen subjects gave informed consent in accordance with a protocol approved by the Georgia Institute of Technology Institutional Review Board prior to participating in the study. Eight people with trans-tibial amputation (5 male, 76.6k±12.3kg, intact leg length = 88.1±5.9cm) and eight control subjects (5 male, 77.4k±11.3kg, intact leg length = 89.1±6.1cm), who were matched by gender, leg length and body weight, took part in this study. Control subjects showed no significant differences from people with trans-tibial amputation in either body mass (mean difference = 0.75kg, p = 0.60) or leg length (mean difference = 1.0cm, p = 0.35). The cause of amputation was traumatic (7 subjects) or congenital (1 subject), and subjects were excluded if they had limited motion in intact joints or were unable to walk for 15 minutes without additional walking aid. People with trans-tibial amputation wore their own, custom-fit prostheses with energy storage and return feet, which they used as their primary prosthesis for at least 6 months prior to the study.

### Experimental Protocol

Because subjects had differing fitness levels, we first determined their preferred walking speeds (PWS). After allowing participants 2 minutes of acclimation to treadmill walking, we determined PWS by asking them to walk on the treadmill at different speeds. Participants started at 0.9m/s and belt speeds increased by 0.1-m/s increments. At each speed, the participant had 30 seconds to indicate if the speed was “too fast,” “too slow” or “comfortable.” If subjects indicated that two consecutive speeds were comfortable, they were asked if the second speed was more comfortable than the first. After increasing to a speed that the subject deemed “too fast,” we repeated the procedure with belt speeds decreasing in 0.1-m/s increments. PWS was determined to be the speed that subjects found comfortable when speeds were both increasing and decreasing. This protocol was similar to a longer, previously used protocol for determining self-selected walking speed (Amorim et al., 2009). Control subjects walked at the PWS of their matched person with trans-tibial amputation, allowing for valid comparisons of mechanical work and other variables. Belt speeds for the rest of the experiment were determined based on PWS.

The beginning of the experimental protocol was similar to a prior study (Selgrade et al., 2017a); subjects completed a baseline trial with both belts moving at 75%PWS (slow baseline 1) followed by a trial with both belts moving at 150%PWS (fast baseline) and a second slow baseline trial (Figure 1). Next people with trans-tibial amputation walked with their prosthetic legs on the slow belt, and the fast belt gradually sped up to 150%PWS from 75%PWS rather than introducing this acceleration suddenly. We chose gradual onset for the split-belt condition, because people with trans-tibial amputation have deficits in standing balance (Seth and Lamberg, 2017), greater ranges of whole body angular momentum during walking (Silverman and Neptune, 2011), and increased risk of and fear of falling (Miller et al., 2001). They also spend less time in stance on their prosthetic leg than on their intact leg (Engsberg et al., 1993, Breakey, 1976), likely because they are more comfortable balancing on the intact leg, which can generate ankle moments to maintain balance when perturbed (Curtze et al., 2012). However, sudden onset split-belt walking presents a greater challenge to both sagittal and frontal plane balance on the slow leg than gradually speeding up the fast belt (Sawers et al., 2013, Sawers and Hahn, 2013).

**Figure 1.**
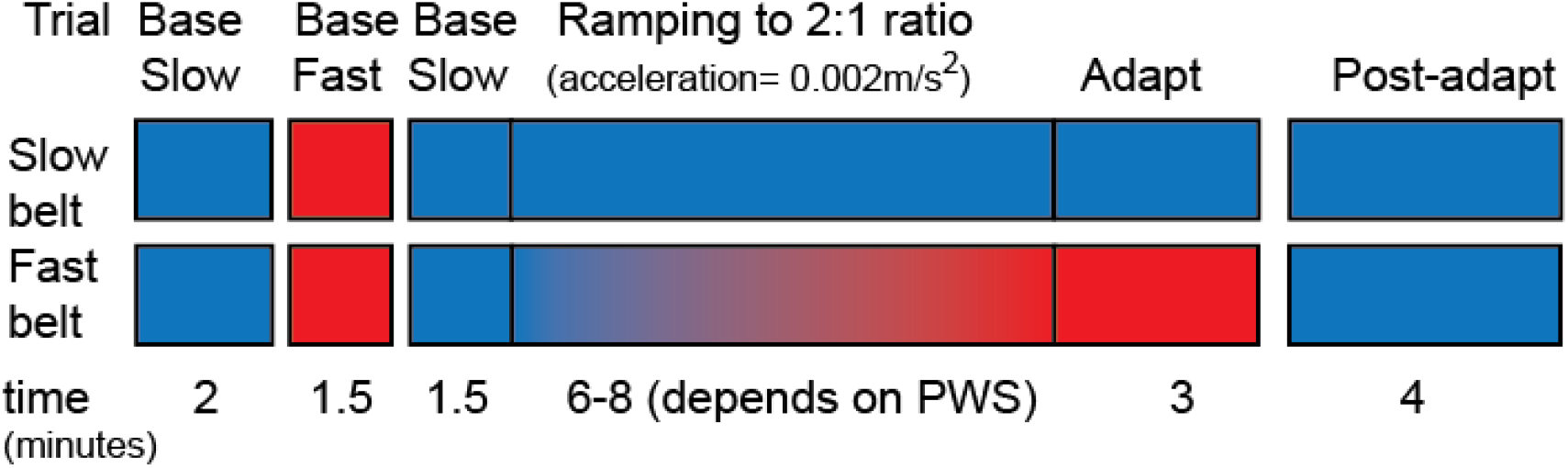
Experimental Protocol. Subjects completed a slow baseline trials at 75%PWS (blue) and a fast baseline trial at 150%PWS (red). Immediately after an additional 1.5 minutes at 75% PWS, the speed of the fast belt was slowly accelerated to 150%PWS.

Similarly to subjects with trans-tibial amputation, intact control subjects walked with their non-dominant leg on the slow belt. The fast belt accelerated at 0.002m/s^2^, a rate at which no participants were able to notice the belt accelerating. After the fast belt reached 150%PWS, there was a 3-minute period where both belts moved at a constant speed and a 2:1 belt speed ratio.

After resting for 3-5 minutes during which they were not permitted to walk, participants completed a 4-minute trial with both belts moving at 75%PWS. This post-adaptation trial was introduced suddenly to allow for detection of an aftereffect without changing belt speed as a potential confounding factor.

To prevent falls, participants wore a safety harness that did not support body weight for all trials. In addition, a mirror in front of the subjects allowed them to see their medial-lateral foot placement. This mirror helped participants avoid stepping on the contralateral belt and stumbling while still maintaining forward gaze and allowed them to step onto the treadmill and let go of the safety bar in front of it quickly during post-adaptation. Post-adaptation trials began when participants let go of the bar, which was within the first 2-5 steps on the treadmill.

### Data Collection and Processing

We collected data in 2-3 minute increments in all experimental conditions. For the adaptation and post-adaptation conditions, 30-second gaps between each increment allowed for the next trial to be set up in the computer system, which only reliably collected a maximum of 2-3 minutes of data at once. For every subject, however, we collected the first 2 minutes and last 30 seconds of belt acceleration and the entire 3-minute period when belts were at a constant 2:1 speed ratio. Similarly, we collected the first 2 minutes and last 90 seconds of post-adaptation.

Because the acceleration of the belt made it difficult to tell if changes in step length, force and other variables were due to changing belt speed or changes in motor control, we focused our analysis primarily on times when belt speed was not changing – baseline, post-adaptation and the last 3 minutes of adaptation. We collected kinematic data using a six-camera motion analysis system (120Hz, VICON Motion Systems, Oxford, UK) and retroreflective markers placed on the anterior posterior iliac spine, posterior superior iliac spine, greater trochanter, thigh, knee, shank, lateral malleolus, heel and second metatarsophalangeal joint of each leg. For people with trans-tibial amputation, markers were placed on the prosthesis at the same locations as on the contralateral, intact leg, as has been done previously (Silverman et al., 2008). We measured GRFs for each leg with mechanically isolated force plates beneath each treadmill (1080Hz, AMTI, Watertown, MA, USA) and processed marker position and GRF data with a 4th order Butterworth filter with a 10Hz cutoff frequency.

Custom-written Matlab programs (MathWorks, Natick, MA) calculated peak vertical and anterior-posterior GRF, negative work and step length symmetry using the same methods as prior work (Selgrade et al., 2017a, Selgrade et al., 2017b). To find step length symmetry (SLS), we calculated the normalized difference between fast-leading and slow-leading step length (SL; Eq 1) (Reisman et al., 2005).

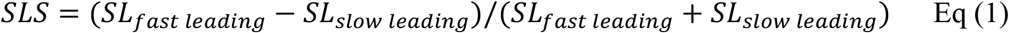

To characterize split-belt adaptation in people with trans-tibial amputation compared to previous studies of healthy subjects and other clinical populations, we calculated other temporal variables. We found double support time as the difference between the times of leading leg heel strike, when the vertical GRF of that leg exceeded 32N, and trailing leg toe off, when vertical GRF decreased below 32N. We also calculated stance time on each leg as the time between ipsilateral heel strike and toe off. In order to find collisional energy loss during braking (W_loss_), we took the negative portion of the integrated dot product of GRF and center of mass velocity (v_com_), using an integration constant based on belt speed (Eq 2).

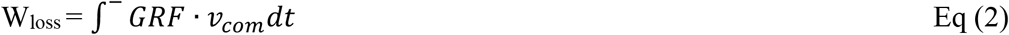

Integration constants were determined based on an average vertical center of mass velocity of zero and an average anterior-posterior velocity equal to treadmill belt speed.

#### Statistical Analysis

We performed statistical analyses with SPSS (IBM, Armonk, NY, USA), defining late adaptation as the averages of the last 5 steps of adaptation. We defined early and late post-adaptation as the first 5 steps and the last 5 steps of post-adaptation, respectively. To analyze step length symmetry, and double support time, we compared slow baseline and early and late post-adaptation using repeated measures ANOVA (rmANOVA) followed by post-hoc, pairwise comparisons using Bonferroni corrections if a significant main effect was found. We compared these three trials, because they had identical conditions with both belts moving at 75% PWS, allowing for a fair comparison that was not affected by walking speed, which can by itself alter gait asymmetries in people with amputations (Nolan et al. 2003). For single limb variables (stance time), we performed 3 rmANOVAs. One rmANOVA compared the fast leg in trials when it moved at 150% PWS (fast baseline, late adaptation); another compared the fast leg when it moved at 75%PWS (second slow baseline, early post-adaptation, late post-adaptation); and the third compared GRFs in all trials but fast baseline. To determine if subjects had baseline step length asymmetries, we found averages across strides in each trial for every subject, and then compared the subject averages to zero using Student’s t-tests for each baseline trial. All statistical tests had an alpha level of 0.05. P-values below 0.05 indicated significant differences while p-values between 0.05 and 0.10 indicated that differences between two groups approached statistical significance.

## Results

As expected for gradual adaptation to split-belt treadmill walking, subjects were able to maintain generally consistent level of symmetry with each step during the adaptation process. Only upon sudden exposure to the tied belt condition in the post-adaptation trial did we observe a dramatic change in inter-limb symmetry. People with trans-tibial amputation and controls both showed clear aftereffects in step length symmetry, taking longer steps with the slow (intact) leg leading than with the fast (prosthetic) leg leading in early post-adaptation. This step length asymmetry in early post-adaptation was significantly higher than slow baseline for controls (Figure 2A; p=0.002), and people with trans-tibial amputation (Figure 2B; p=0.015). For each group, this difference washed out by late post-adaptation, at which point step lengths were significantly more symmetric than early post-adaptation for controls (p=0.002) and people with trans-tibial amputation (p=0.006). The pattern of step length symmetry as the fast belt was accelerating, shows that, for both groups of subjects, step length asymmetry became more negative (i.e. asymmetric with longer steps when the fast leg is leading) in the first 100 strides but stayed relatively constant for the remainder of the adaptation period, even while the belt was still accelerating. This pattern of slight changes during belt acceleration was generally maintained in other variables. In the fast baseline trials, people with trans-tibial amputation had an average step length asymmetry that was significantly less than zero (p=0.0467).

**Figure 2.**
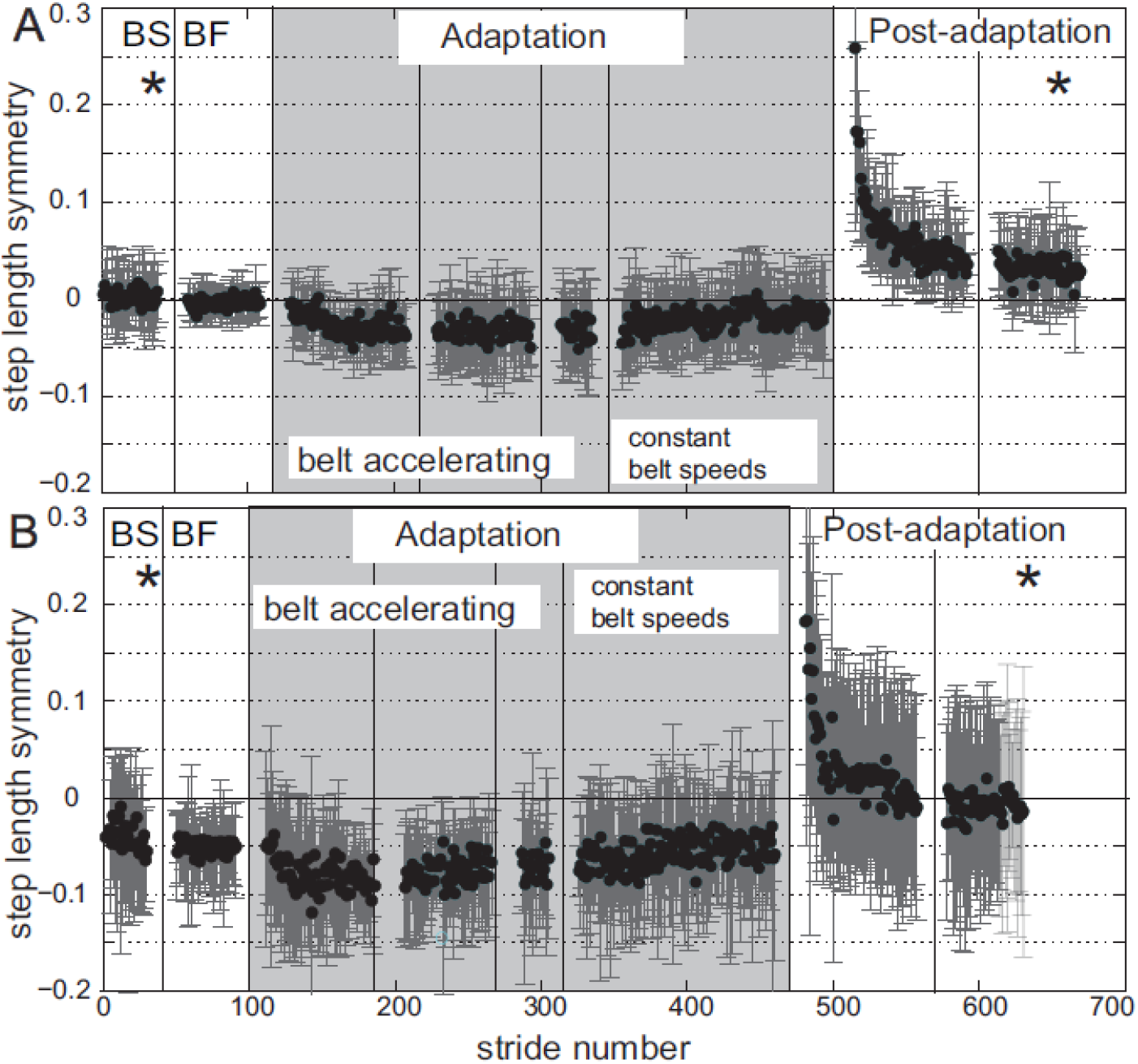
Step length symmetry averaged across intact control subjects (A) and people with trans-tibial amputation (B). BS and BF are slow baseline (belts at 75% preferred walking speed) and fast baseline (belts at 150% preferred walking speed), respectively. Asterisks indicate significant differences from early post-adaptation. In late adaptation, only the constant belt speed portion of adaptation is shown. For descriptive purposes, the portion of the split-belt adaptation trial when the belts are accelerating is shown. Error bars denote standard deviation.

There were several significant changes in GRF from baseline to post-adaptation. People with trans-tibial amputation had significantly lower peak propulsive GRF from the fast (intact) leg and higher peak propulsive GRF from the slow (prosthetic) leg during early post-adaptation compared to slow baseline and late post-adaptation (Figure 3B, p≤0.03 for all). This means that, in people with trans-tibial amputation, the intact limb produced less force than the prosthetic leg in the aftereffect, reversing their baseline asymmetry. Controls showed a similar pattern, with peak propulsive GRF from the fast leg were significantly less in early post-adaptation than in slow baseline (p=0.010) or late post-adaptation (p=0.007). Although there was a significant main effect of condition on slow leg peak propulsive GRF (p=0.048), pairwise comparisons showed differences in slow leg propulsive GRF for controls were not significant for either leg (p>0.20; Figure 3A). In early post-adaptation, people with trans-tibial amputation (p=0.014) and controls (p=0.035) both had significantly higher fast leg peak braking GRF than in the slow baseline trial (Figure 4). Controls significantly decreased slow leg peak braking GRF in early post-adaptation (p<0.001), but people with trans-tibial amputation did not show any post hoc differences in slow leg braking GRF between trials (p=0.130), although there was a significant main effect (p=0.017).

**Figure 3.**
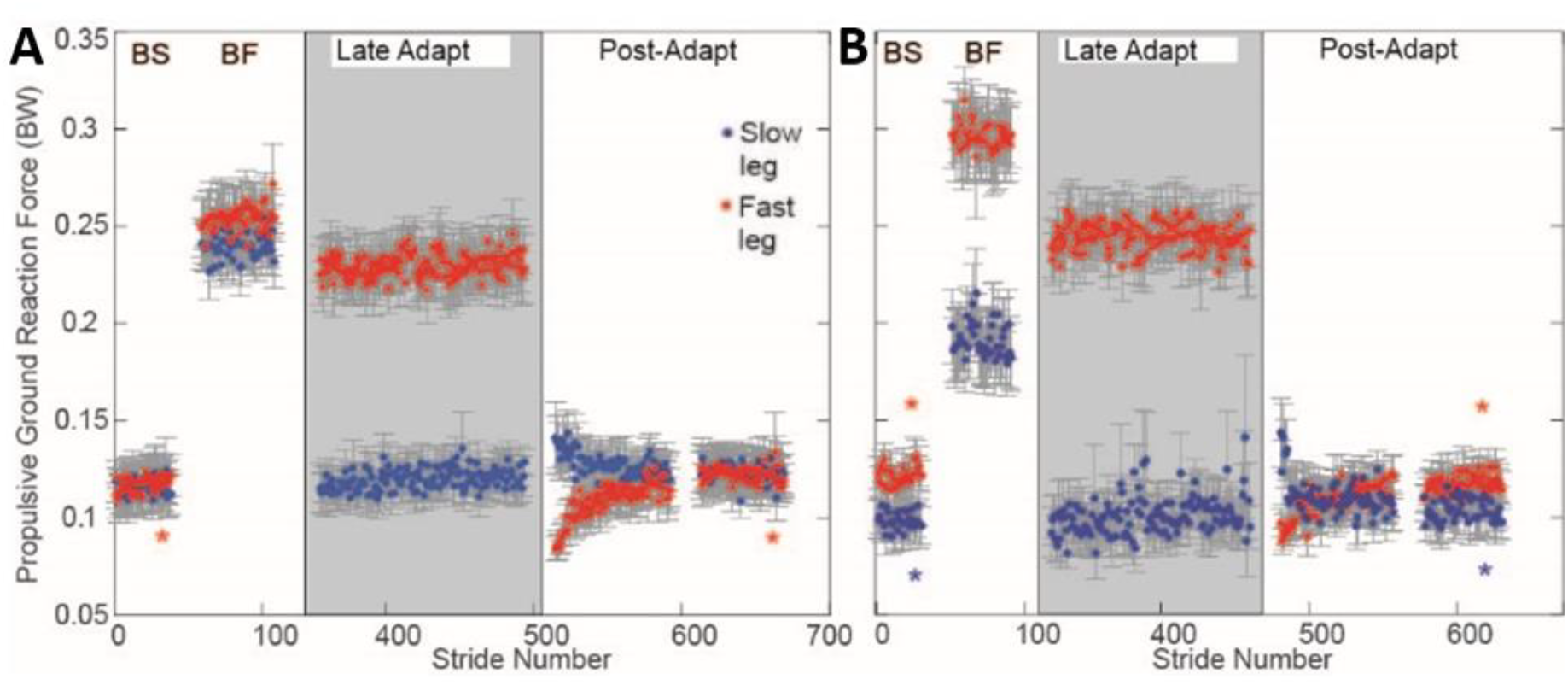
Peak propulsive GRF for control subjects (A) and people with trans-tibial amputation (B). Red * indicate significant difference from early post-adaptation for fast leg. Blue * indicate significant difference from early post-adaptation for slow leg. Circles and error bars denote mean and standard error across subjects, respectively. In late adaptation, only the constant belt split-belt speed (2:1 speed ratio between belts) portion of adaptation is shown.

**Figure 4.**
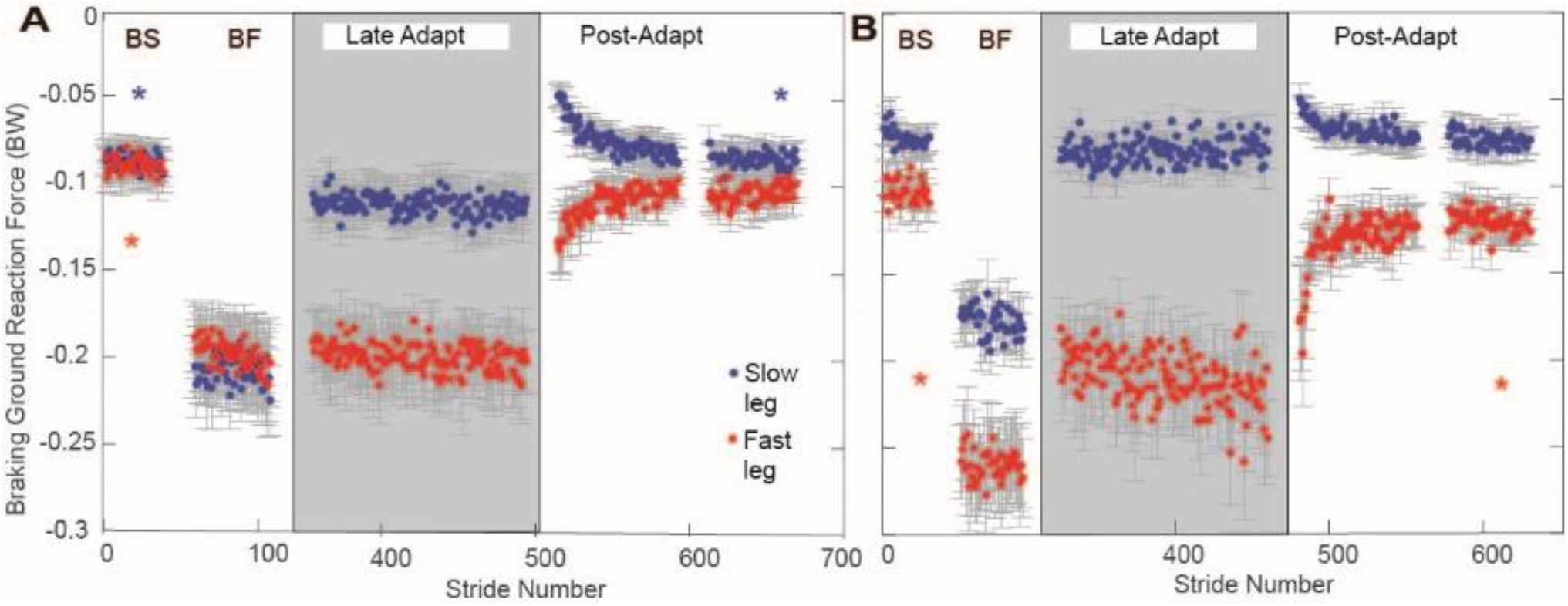
Peak braking GRF for control subjects (A) and people with trans-tibial amputation (B) – Red * indicate significant difference from early post-adaptation for fast leg. Blue * indicate significant difference from early post-adaptation for slow leg. Circles and error bars denote mean and standard error across subjects, respectively. In late adaptation, only the constant belt split-belt speed (2:1 speed ratio between belts) portion of adaptation is shown.

Collisional energy loss showed similar trends to braking GRFs. For people with trans-tibial amputation, collisional energy loss at the fast leading leg was significantly higher in early post-adaptation than slow baseline (p=0.021) and approached being significantly higher in early post-adaptation than late post-adaptation (p=0.059). For controls, collisional energy loss at the fast leading leg approached being significantly higher in early post-adaptation than slow baseline (p=0.082).

During early post-adaptation, all subjects spent less time in double support when the fast leg was leading (Figure 5). Controls had shorter fast-leading double support times in early post-adaptation than in slow baseline (p<0.001; Figure 5A) and late post-adaptation (p=0.012). People with trans-tibial amputation also had shorter fast-leading double support times in early post-adaptation than in slow baseline (p=0.003; Figure 5B) and late post-adaptation (p=0.026). For control subjects, slow-leading double support time was significantly greater in early post-adaptation than in slow baseline (p=0.003) and late post-adaptation (p=0.004; Figure 6A), but slow-leading double support time showed no significant differences between trials for people with trans-tibial amputation (p>0.21, Figure 6B). In late adaptation, fast (intact) leg total stance time was significantly shorter than in fast baseline (data not shown; p<0.001 for controls, p=0.021 for people with trans-tibial amputation). Slow (prosthetic) leg total stance time was significantly longer in late adaptation than in all other trials (p≤0.016 for controls, p≤0.030 for people with trans-tibial amputation).

**Figure 5.**
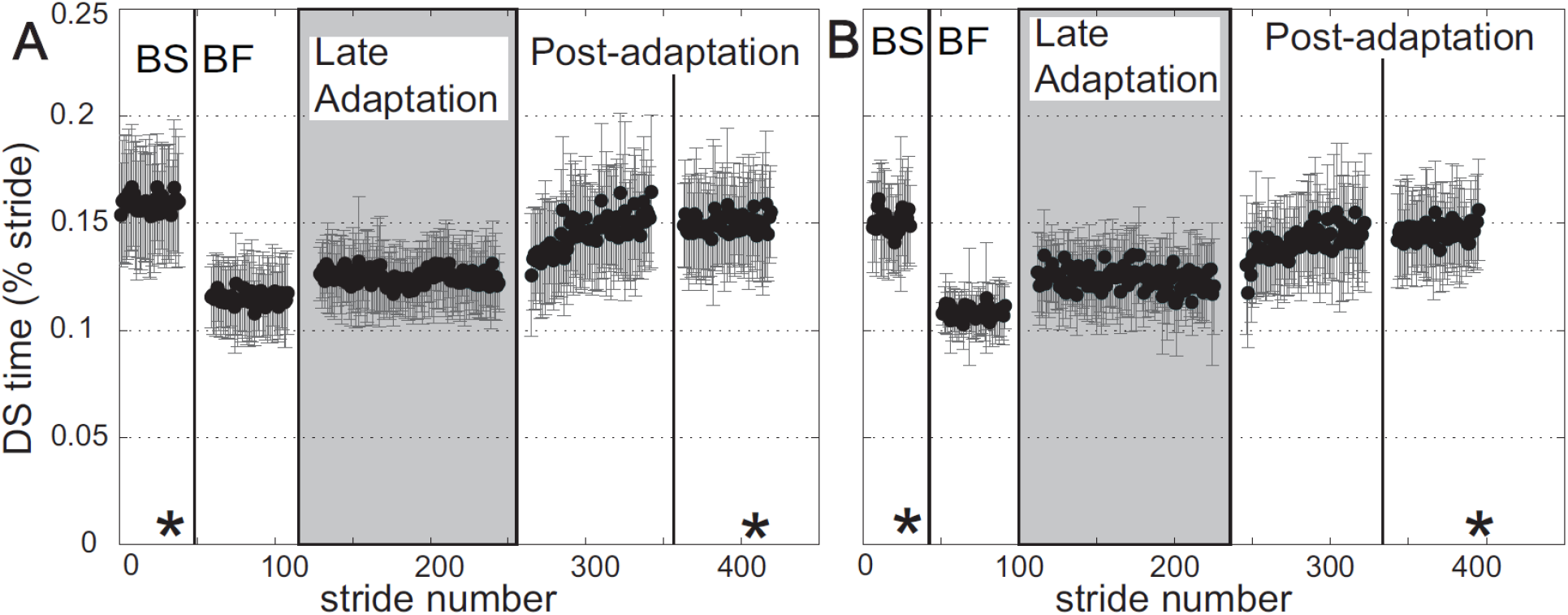
Double support time when the fast leg is leading for control subjects (A) and people with trans-tibial amputation (B). Asterisks indicate significant differences from early post-adaptation. Circles and error bars denote mean and standard deviation across subjects, respectively. BS and BF are slow and fast baseline trials (tied belt) and late adaptation is the last 3 minutes of adaptation, when belt speeds are at a constant 2:1 ratio.

**Figure 6.**
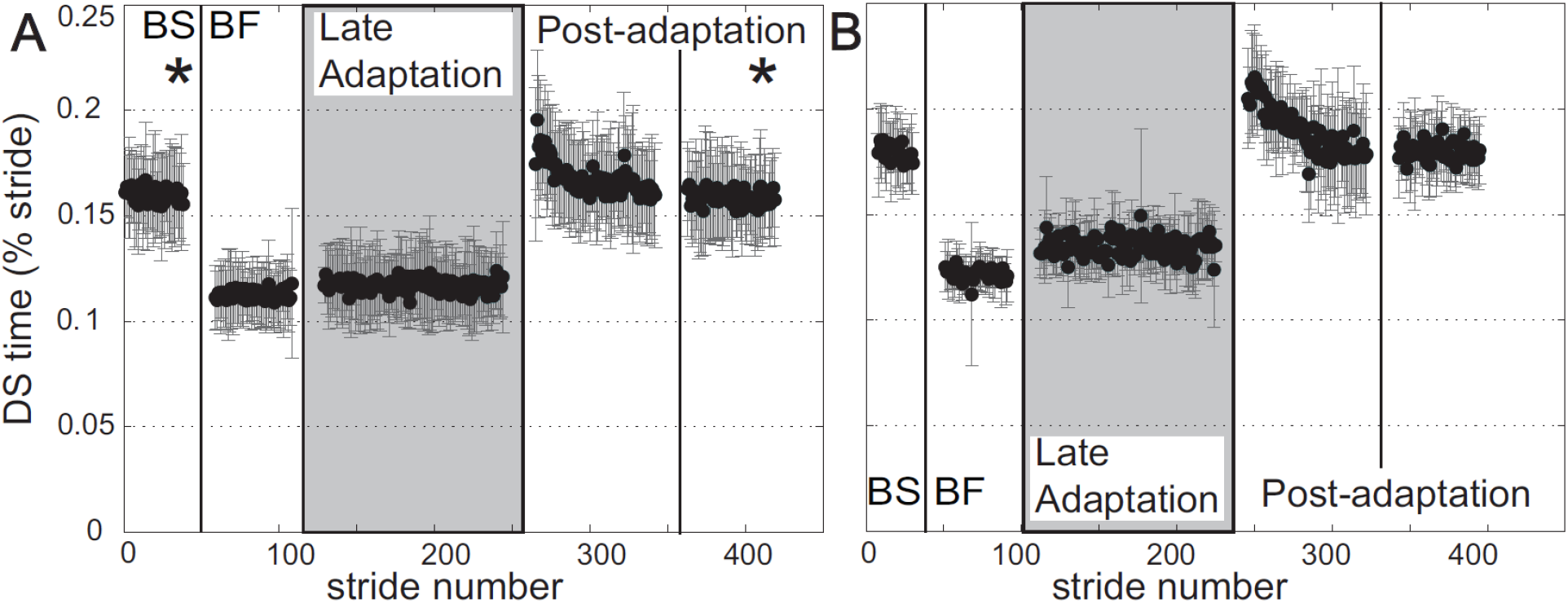
Double support time when the slow leg is leading for control subjects (A) and people with trans-tibial amputation (B). Asterisks indicate significant differences from early post-adaptation. Circles and error bars denote mean and standard deviation across subjects, respectively. BS and BF are slow and fast baseline trials (tied belt) and late adaptation is the last 3 minutes of adaptation, when belt speed are at a constant 2:1 ratio.

## Discussion

### Kinetic Adaptations to Split-belt Walking

The hypothesis that, in early post-adaptation, subjects would display aftereffects in peak braking GRF was supported in healthy control subjects and people with trans-tibial amputation. Post-hoc, pairwise comparisons here are consistent with previous work on sudden split-belt walking (Ogawa et al., 2014) and suggest that people with intact legs use predictive control to modulate peak braking GRF in early stance, resulting in an aftereffect in which slow (intact) leg braking GRFs are lower in magnitude and fast leg braking GRFs are higher in magnitude. This control strategy could relate as much to the timing of peak braking GRF as to its direction.

Shortly after heel strike, there is too little time for ipsilateral reflexes from that stance phase to reach the spinal cord and alter motor commands to change the concurrent braking GRF (Dietz, et al, 1979, Toft et al., 1991). Therefore, the motor control system seems to rely on a predictive controller that is updated by sensory feedback from many previous steps.

The hypothesis that peak propulsive GRF would indicate reactive control by failing to show aftereffects was not supported. People with trans-tibial amputation had aftereffects in peak propulsive GRF, increasing propulsion from the slow (prosthetic) leg and decreasing fast leg propulsion in early post-adaptation. Controls had significantly lower fast leg propulsive GRF in early post-adaptation. These findings indicate predictive control of propulsive GRF in people with trans-tibial amputation and controls. This is surprising compared to prior results; Ogawa and colleagues found a significant aftereffect in propulsive GRF of only the slow leg (2014). Given the similarity between amputees and healthy control subjects in other split-belt adaptation studies (Selgrade et al. 2017a, Darter et al. 2017, Kline et al., 2019), we expected their limb forces to adapt similarly as well. However, more recent studies have shown aftereffects in propulsive forces after inclined split-belt walking (Sombric et al., 2019, Sombric and Torres-Oviedo, 2020) and in people with multiple sclerosis (Hagen et al., 2024) after split-belt walking. In each of these cases, there was greater propulsive demand during split-belt walking due either to the inclined treadmill or to participants having multiple sclerosis, which limits propulsion (Cofre Lizama et al., 2016). In contrast, studies that failed to show aftereffects in GRF used a slow belt speed of 0.5 m/s fast belt speed of 1 m/s (Ogawa et al., 2014, Mawase et al., 2013), both of which are slower than an average person’s preferred walking speed. In the present study, the fast belt speed was greater than 1m/s for all subjects. It is possible that prior work could not detect propulsive force aftereffects given the slow belt speeds and resulting limited need for propulsion. When more propulsion is demanded, it appears that people use predictive control of propulsive force. Because step length is altered by propulsive force, predictive control of propulsive force is also consistent with the adaptation of step length symmetry observed here and in most spilt-belt studies.

Alternatively, because the current study is the first to examine forces during gradual split-belt adaptation, it is possible that gradual exposure to the split-belt condition caused the discrepancy between the two studies. When visuomotor perturbations are introduced gradually during reaching, aftereffects are larger than if such perturbations are introduced suddenly (Kagerer et al., 2007). Perhaps gradual introduction of split-belt walking allows subjects to more robustly adapt their limb forces, producing larger aftereffects than previous, sudden onset studies. In prior studies, gradual and sudden onset split-belt walking show similar washout of step length symmetry (Torres-Oviedo and Bastian, 2012, Roemmich and Bastian, 2015), but kinetic adaptations to gradual onset split-belt walking may not show the same patterns as kinematics. Although the current results indicate predictive control of fast leg propulsive GRF for both groups and slow leg propulsive GRF for people with trans-tibial amputation, differences in control subjects’ slow leg propulsive GRF were not significant, possibly because each group only had eight individuals. Future work should compare kinetic responses to gradual onset and sudden onset split-belt walking with more subjects to determine if predictive control of propulsive GRF is related to the onset of the split-belt condition.

### Inter-limb coordination is similar in people with trans-tibial amputation and controls

The hypothesis that people with trans-tibial amputation would demonstrate an aftereffect in inter-limb coordination that opposed their baseline asymmetry was supported. People with trans-tibial amputation actually overcorrected their baseline step length asymmetry, so the aftereffect was even larger than we initially expected. In early post-adaptation, people with trans-tibial amputation took longer steps with the intact leg leading than with the prosthesis leading, and this asymmetry was opposite and larger in magnitude than their baseline asymmetry towards taking longer steps with the prosthesis leading. Darter and colleagues found that, when exposed to suddenly introduced split-belt walking with one belt moving three times faster than the other, people with trans-tibial amputation adapted their step length in a manner consistent with predictive control (2017). Our results support and extend Darter and colleagues’ findings to gradual onset split-belt walking at a 2:1 belt speed ratio, and are consistent with split-belt walking aftereffects previously found in able-bodied subjects (Reisman et al., 2005, Huynh et al., 2014). Furthermore, this study is the first to demonstrate that people with trans-tibial amputation, like control subjects, display aftereffects in double support time symmetry. The aftereffect and subsequent de-adaptation back to symmetry in step length and double support time provide robust evidence that people with trans-tibial amputation use predictive control inter-leg coordination for both spatial and temporal parameters. Given that, under several different split-belt conditions, adaptation of inter-leg parameters is similar between people with trans-tibial amputation and intact individuals, it is reasonable to conclude that feedback from the distal limb is not necessary for predictive control of inter-leg coordination during locomotor adaptation.

Neither people with trans-tibial amputation nor controls exhibited an aftereffect in slow (prosthetic) leg stance time. Both groups had slow leg stance times that were significantly different in late adaptation from baseline, but stance time immediately returned to slow baseline levels in post-adaptation, indicating that stance time is likely controlled reactively based on sensory feedback. This is consistent with prior studies of people with trans-tibial amputation (Darter et al., 2017) and healthy individuals (Reisman et al., 2005). Given that amputation compromises sensory feedback from the amputated leg during stance, reactive accommodation of stance time must not be reliant on feedback from the foot or afferent sensors in muscles crossing the ankle. Rather, people with trans-tibial amputation likely use sensory feedback from proximal muscles and joints. This finding is consistent with control of cat locomotion, in which feedback from hip afferents trigger the spinally mediated stance-swing transition (Hiebert et al., 1996). This result does not necessarily mean that feedback from the ankle and foot has no role in the stance-swing transition, but it suggests that this transition can be controlled reactively without feedback from the lower leg.

### Clinical Implications

The clinical goal of split-belt adaptation research is to create an aftereffect that corrects a patient’s baseline asymmetry. After repeated bouts of split-belt adaptation, this aftereffect could become more enduring, as has been shown with step length symmetry in a portion of stroke survivors (Reisman et al., 2013). In the present study, people with trans-tibial amputation not only correct but overcorrect their baseline step length asymmetries. Asymmetry involving longer steps with the intact leg leading is not clinically desirable, and aftereffects that overcorrect baseline asymmetry to produce the opposite asymmetry washout more quickly than smaller aftereffects (Tyrell et al., 2015). Therefore, long-term studies that seek to induce kinematic symmetry in people with trans-tibial amputation may benefit from using smaller belt speed ratios based on each individual’s baseline asymmetry to induce smaller, but longer lasting, step length aftereffects.

Aftereffects in GRF have more complex clinical implications for people with trans-tibial amputation. In early post-adaptation, people with trans-tibial amputation increased propulsive GRF from the slow (prosthetic) leg and decreased propulsive GRF from the intact leg. If these aftereffects could transfer to overground walking after repeated split-belt exposures, that would accomplish the goal of lowering forces experienced by the intact leg, potentially reducing overuse injuries. However, a limitation of this study was that it did not include modeling of loads in the intact side knee or hip, common sites for osteoarthritis after trans-tibial amputation (Royer and Wasilewski, 2006, Baliunas et al., 2002). Recent analyses show that decreasing step length decreases intact limb knee contact forces in those with trans-tibial amputation (Syrett et al., 2025), but such analyses have not been applied to split-belt walking in people with amputation.

Although induced asymmetries that alter GRFs do not always change joint loading (McCain et al., 2023), it is encouraging that, after split-belt walking, healthy individuals exhibit aftereffects including lower average contact forces in the knee of the fast leg (Syrett et al., 2021). This result would translate to the prosthetic leg in the current study. However, it is unclear if people with trans-tibial amputation would adapt similarly to controls or if they would compensate for an adaptation that lowers forces in the knee by increasing forces in the hip, which may not be a desired clinical result. Therefore, more research is necessary to fully understand how split-belt walking affects joint loading in people with trans-tibial amputation.

Peak braking GRF on the intact limb increased in the aftereffect. This increase in braking force was accompanied by an increase in energy lost during intact leg braking, which is already a difficulty with prosthetic gait and would likely cause people with trans-tibial amputation to expend more energy when walking (Houdijk et al., 2009). Moreover, higher braking forces could put more stress on a limb that is already at increased risk of osteoarthritis. Thus, split-belt walking induced potentially beneficial aftereffects during propulsion, but potentially harmful aftereffects during braking, although these conclusions are limited by the small sample size of the current study. Future studies should involve more participants and model changes in joint loading after split-belt walking to better understand how much these limb-level results could affect development of osteoarthritis in people with trans-tibial amputation. Future work could also determine if these aftereffects would become permanent after repeated adaptation and transfer to overground walking.

## Conclusion

Despite lacking a distal limb, people with trans-tibial amputation adapt to split-belt walking similarly to intact control subjects. This study shows for the first time that double support time, and propulsive and braking GRFs in people with trans-tibial amputation adapt in a manner consistent with predictive control. This work also supports the results of prior research by indicating that step length symmetry in those with trans-tibial amputation is subject to predictive control while stance time on each leg is subject to reactive accommodation. These results provide robust evidence that trans-tibial amputation does not interfere with predictive adaptation of ground reaction forces and inter-leg coordination. Additionally, step length symmetry actually overcorrected in early post-adaptation, producing an asymmetry that was opposite the baseline asymmetry and larger in magnitude. Finally, aftereffects in ground reaction forces suggest that gradual onset split-belt adaptation involves predictive control of both braking and propulsion, in contrast to prior results in sudden onset split-belt walking. Future work should consider joint kinetics to ensure that split-belt training does not increase loads in areas already at risk of overuse injury after trans-tibial amputation.

## Data Availability

All data produced in the present study are available upon reasonable request to the authors

## Acknowledgements

We thank Dawit Lee and Parth Patel for their assistance with data processing and Eunse Park, Kevin Hetzendorfer and Sam Kwak for their helpful comments and assistance with data collections. This research was supported by NICHD 5T32HD055180, NINDS 5R01NS069655, NSF ARRA CAREER Award BCS-0847325 and an NSF Graduate Research Fellowship.

